# Evaluating the Effect of Prebiotics on the Gut Microbiome Profile and β-cell Function in Youth with Newly-Diagnosed Type 1 Diabetes: Protocol of a Randomized Controlled Trial

**DOI:** 10.1101/2020.10.05.20207142

**Authors:** Heba M. Ismail, Maria Spall, Carmella Evans-Molina, Linda A. DiMeglio

**Author notes:** Corresponding author: Heba M. Ismail, MB BCh, MSc, PhD, 635 Barnhill Drive | 2053, Indianapolis, IN 46202.

## Abstract

**Introduction:** Data show that disturbances in the gut microbiota play a role in glucose homeostasis, type 1 diabetes (T1D) risk and progression. The prebiotic high amylose maize starch (HAMS) alters the gut microbiome profile and metabolites favorably with an increase in bacteria producing short chain fatty acids (SCFAs) that have significant anti-inflammatory effects. HAMS also improves glycemia, insulin sensitivity and secretion in healthy non-diabetic adults. Additionally, a recent study testing an acetylated and butyrylated form of HAMS (HAMS-AB) that further increases SCFA production prevented T1D in a rodent model without adverse safety effects. The overall objective of this human study will be to assess how daily HAMS-AB consumption impacts the gut microbiome profile, SCFA production, β-cell heath, function and glycemia as well as immune responses in newly-diagnosed T1D youth.

**Methods and Analysis:** We hypothesize that HAMS-AB intake will improve the gut microbiome profile, increase SCFA production, improve β-cell health, function and glycemia as well as modulate the immune system. We describe here a pilot, randomized crossover trial of HAMS-AB in 12 newly-diagnosed T1D youth with residual β-cell function. In **Aim 1**, we will determine the effect of HAMS-AB on the gut microbiome profile and SCFA production; in **Aim 2**, we will determine the effect of HAMS-AB on β-cell health, function and glycemia; and in **Aim 3**, we will determine the peripheral blood effect of HAMS-AB on frequency, phenotype and function of specific T cell markers. We anticipate beneficial effects from a simple, inexpensive, and safe dietary approach.

**Ethics and Dissemination:** The Institutional Review Board at Indiana University approved the study protocol. The findings of this trial will be submitted to a peer-reviewed pediatric journal. Abstracts will be submitted to relevant national and international conferences.

**Trial registration number:** NCT04114357; Pre-results.

**Article Summary:** *Strengths and limitations of this study:*

- The study design (randomized controlled trial, RCT) is the most robust methodology to assess the effectiveness of therapeutic interventions.
- The findings of this RCT, whether positive or negative, will contribute to the formulation of further recommendations on the use of *high amylose maize starch that has been acetylated and butyrylated* for improving beta-cell function in children with newly diagnosed type 1 diabetes (T1D).

## Introduction

Compared to healthy individuals, people with type 1 diabetes (T1D) demonstrate gut dysbiosis (microbial imbalance) with higher gram-negative to gram-positive gut bacterial ratios, creating a pro-inflammatory milieu, and fewer bifidobacteria, which are an important microbial population associated with health and anti-inflammatory effects [1-9]. Further, the gut microbiome composition of children with T1D shows increased virulence factors, phage, prophage and motility genes, and T1D children display a lower count of bacteria producing butyrate, a short chain fatty acid (SCFA) with anti-inflammatory actions [10-13].

High amylose maize starch (HAMS) is a dietary fiber and prebiotic that provides a novel approach to mitigating diabetes through altering the gut microbiome. HAMS is a resistant corn starch type 2 (RS2) derived from high amylose maize starch and containing approximately 70% amylose. RS2 is not digested in the small intestine and acts as a dietary fiber. HAMS consumption shifts the gut microbiome profile towards dietary fiber fermenters producing abundant SCFAs (specifically acetate and butyrate) [14]. Further, HAMS that is acetylated and butyrylated (HAMS-AB) releases large amounts of beneficial SCFAs after colonic bacterial fermentation. Indeed, recent T1D non-obese diabetic (NOD) mouse studies [14] have shown that HAMS-AB feeding shifted the gut microbiome profile towards fermenters, resulting in higher blood and fecal concentrations of acetate and butyrate (both beneficial SCFAs). These changes were associated with significantly lower rates of T1D progression, lower autoreactive T cells, increased regulatory T cells, and decreased islet autoimmunity. Limited non-diabetic adult human data indicate HAMS lowers post-prandial glucose production and improves insulin sensitivity [15, 16].

The exact link between dysbiosis and immune cell activation remains unclear. Mucosal-associated invariant T (MAIT) cells are innate-like (innate immune responses provide first line of defense against non-self-pathogens and typically innate-like cells are found at barrier tissue like the gut) T-cells whose development is dependent on the host microbiome. MAIT cells are altered (showing evidence of decreased frequency and function) in children with T1D [17] and likely play a key role in maintaining gut integrity by controlling the autoimmune response to β-cells, thereby potentially providing a link between the gut microbiome changes and autoimmunity. A study by Rouxel et al [17] found altered MAIT cell populations (lower overall count along with functional alteration upon stimulation in vitro) in both NOD mice and persons with T1D. Further, they demonstrated that gut mucosal MAIT cells in the mice were found to produce protective cytokines essential for intestinal homeostasis, namely IL-17A and IL-22. Moreover, MAIT cell genetic ablation in MR1 knockout mice (MR1 is required for the thymic development of MAIT cells) was associated with a loss of gut integrity. Indeed, the data from this study support a model in which MAIT cell deficiency aggravates alterations to the gut mucosa and thereby promotes the development of T1D. Overall, these data suggest that MAIT cells might be a nexus between the gut microbiome and gut mucosa, inflammation and T1D. Given the above, we **hypothesize** that HAMS-AB consumption will alter the gut microbiome profile in humans with T1D and increase SCFA production and that these changes will be associated with improved β-cell health (as measured by markers of β-cell stress), function and overall glycemia as well as improved number and function of MAIT cells.

## Methods and Analysis

### Overall Experimental Design

This is a single-center pilot randomized crossover trial using HAMS-AB in 12 youth with newly-diagnosed T1D. We will use state-of-the-art markers to profile the gut microbiome and asses β-cell health, function and glycemia as well as describe frequency, phenotype and function of MAIT cells. **Figure 1** shows a schematic for the overall study design.

**Figure 1:**
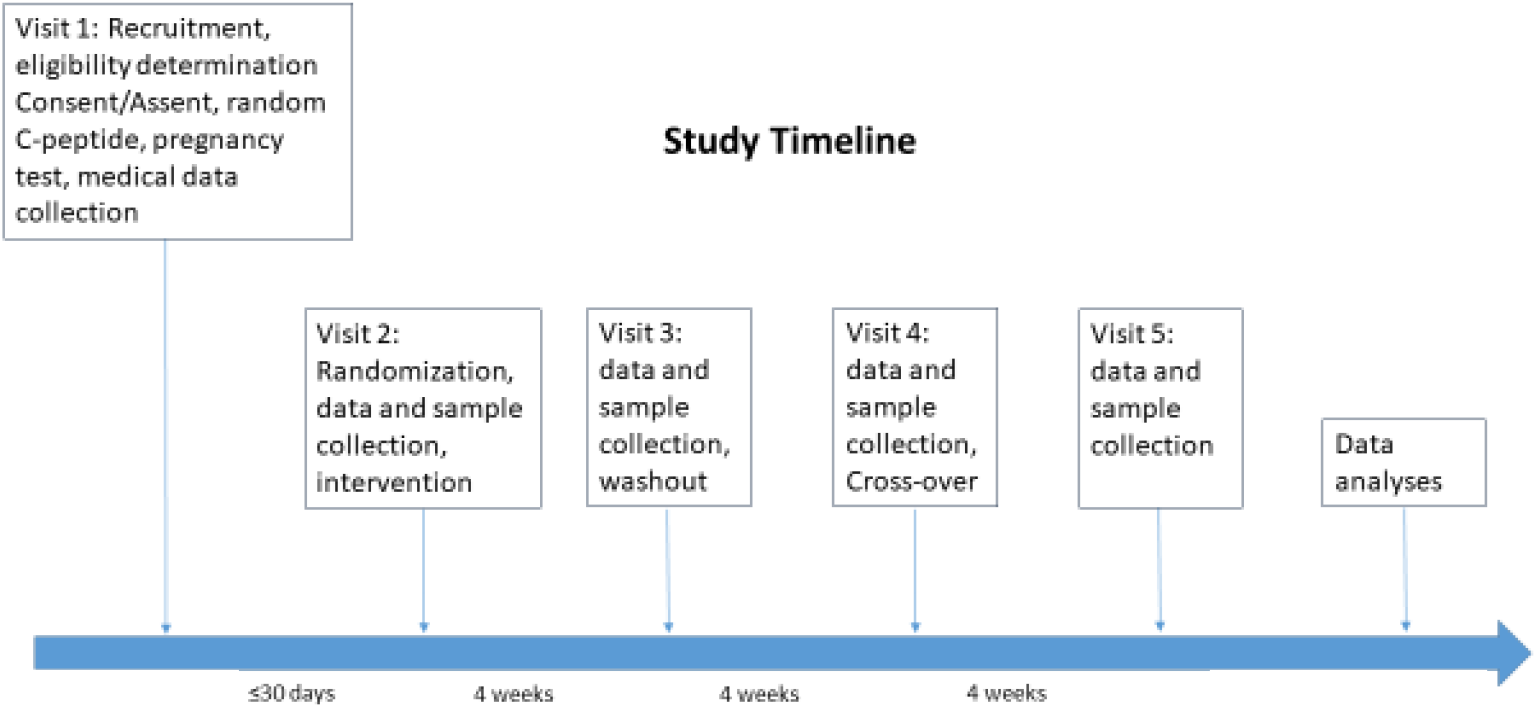
Study Design.

### Participants

This pilot study has been approved by the Indiana University Institutional Review Board (IRB protocol #1907172784). It has also been registered with <u>ClinicalTrials.gov</u>. Participants are being recruited from the diabetes clinics at Riley Hospital for Children at Indiana University Health. Advertisements and flyers have also been posted, including on social media sites. Up to 24 subjects will be screened to find 12 adequate subjects eligible for enrollment. The plan is to recruit 24 patients with the goal of having 50% eligible and enrolled.

Consent from legally authorized representatives/assent from participants will be obtained after members of the study team explain the study in detail, including the need for participants to use a reliable form of barrier contraception during the study, if sexually active. A medical interview will then be performed with participants and their parents to determine further eligibility for the study, in addition to meeting the inclusion criteria listed below.

Subjects will be randomly assigned (6 in each arm) to either HAMS-AB plus the standard recommended diabetic diet (see below) or standard diabetic diet alone (control) for each of the 4 weeks of treatment period. There will be a 4 week washout period (time between the two treatment periods to allow for the effect of either the prebiotic and/or diabetic diet to washout). After the washout period, individuals will cross over to the other arm. Dietary guidance will follow the International Society for Pediatric and Adolescent Diabetes guidelines (ISPAD) [18]. In addition to randomization and following diabetic diet guidelines, the cross-over design minimizes the influence of several confounding factors that could influence the effect of HAMS-AB such as dietary variables, physical activity, genetics, etc.

Randomization will be done using a computer-based randomization plan performed through RedCap. This plan will be prepared by an individual not involved in the study. As a subject is screened and qualifies, he/she will be assigned in order of screening date on the computer-generated document. One master list will be maintained in a confidential central shared folder for use. Subjects will be replaced on an ongoing list if they drop out.

### Inclusion criteria

Children (ages 12-16 years) with a BMI <85% for age and sex with T1D and within 4-24 months from diagnosis will be recruited. We chose this age range to study since the microbiome profile differs by age [19] and BMI [20], and the progression of T1D differs between children and adults [21]. Additionally, since previous research by members in our group have shown microbiome changes in response to other forms of prebiotic supplementation in healthy adolescents [22, 23], we will study a similar age group with available published data. We will include persons 4-24 months from diagnosis as this represents a “honeymoon period” where children have sufficient measurable C-peptide secretion to accurately assess β-cell function changes. The honeymoon period is the time shortly after T1D diagnosis when the β-cells undergo partial yet temporary recovery and produce enough insulin to reduce insulin needs and aid with glycemic control [24]. Residual insulin secretion will be assessed using a peripheral blood random C-peptide of ≥0.17nmol/L as an eligibility criterion for the study, as has been used in other new onset T1D studies [25, 26]. They will also be asked to consume a sample of the prebiotic mixed with food at the screening visit to ensure that they will tolerate taking it during the study and prior to randomization, **Table 1**.

**Table 1:** Participant Inclusion/Exclusion Criteria.

| Inclusion Criteria | Exclusion Criteria |
| --- | --- |
| • **Children (aged 12-16 years)**<br>• **BMI <85% for age and sex**<br>• **T1D duration of 4-24 months**<br>• **Random C-peptide >0.17nmol/L**<br>• **Able to consume test dose of prebiotic** | • **Monogenic forms of diabetes or type 2 diabetes**<br>• **History of ongoing infection or antibiotic treatment within the past 3 months**<br>• **History of immune-compromise, recurrent infections, steroid intake (inhaled or oral forms) or other immunosuppressant use in the past 6 months**<br>• **History of chronic gastrointestinal disease, including possible or confirmed celiac disease**<br>• **Pregnancy or possible pregnancy**<br>• **Allergy to corn (prebiotic) or milk, soy (present in the mixed meal) or their products**<br>• **Participation in other intervention research trials within prior 3 months**<br>• **Anticipated major change in diabetes management during the** |
|  | **study (e.g., change from injections to insulin pump therapy or new to continuous glucose monitor usage)**<br>• **Consuming a high-fiber or vegetarian diet (consuming three or more servings of high-fiber foods on 4 or more days per week) or any fiber supplements.** |

### Exclusion criteria

Individuals who meet any of the exclusion criteria listed in Table 1 at the time of recruitment will not be enrolled.

### Intervention

HAMS-AB will be consumed in the form of acetylated and butyrylated HYLON^®^ VII (Ingredion Incorprated, Bridgewater, NJ). After fecal/blood collection at their first visit, participants will be randomized to HAMS-AB or no supplement twice daily for four weeks; then undergo a four week washout period. They will then cross-over to the other study arm. HAMS-AB doses will be measured by the Investigational Drug Services at Indiana University and provided in bottles (2 each day) in powder form. Participants will be instructed to mix the prebiotic with food (examples given include oatmeal, applesauce or pudding) and consume it twice daily: once in the morning and then once in the evening. The dose administered will be the one previously described for children; 10g plus 1g per year of age daily [27].

The study coordinator will assess adherence by bottle counting/weighing as well as participant/caregiver interviews. Subjects who miss 20% of prebiotic doses (11 of 56 doses) will be replaced. Each 4□week period will end with an in-person visit to assess for any medical history changes and to collect stool and blood samples for our outcome variables, resulting in a total of 4 study visits in addition to the initial screening visit. Safety and tolerability will be assessed throughout the study. A 3-day log for insulin history will be also collected at all visits and diet recall data will be collected using a validated tool (see below) to assess the diet in the 24 hours prior to stool collection. Children and their caregivers will be instructed and provided with printed materials with guidance on following the recommended diabetic diet at home during both four-week intervention periods, consistent with current recommendations for age [18]. Their diet during the washout period will be self-selected and data will be also collected using the ASA24 at the end of the 4-week period.

### Ongoing Diabetes Management

All study participants will have a treating endocrinologist who is primarily responsible for their diabetes care. In general, the expectation is that all participants will receive at least three injections of insulin daily, including short- and long-acting insulin preparations, or will utilize continuous subcutaneous insulin infusion (CSII, insulin pump). As part of diabetes monitoring during the subjects’ participation, we will encourage participants to check glucose levels at least four times daily if using blood glucose monitor and if not already using a CGM. **At the time of visits the study team** will suggest changes that they believe would improve the glucose control, if necessary. Letters summarizing the study visit will also be sent to the endocrinologist after parental agreement.

### Study procedures

#### Stool Sample Collection and Analysis

Stool samples will be collected at home using a commercially available kit (Zymo feces catcher and RNA/DNA shield fecal collection tubes) within 3 days of presentation to each visit.

- Fecal samples will be obtained for gut microbial community analysis at baseline and prior to each visit during the study period. DNA will be extracted from 50–100mg of fecal material using the FastDNA SPIN Kit for Soil (MP Biochemicals). DNA will be quantified using a NanoDrop 3300 fluorospectrometer (Thermo Scientific). Microbial community composition will be determined by PCR-amplification of the 16S rRNA gene from the DNA extracts, using Illumina MiSeq sequencing (PE250).
- Additional *metagenomic sequencing* will be performed [28, 29]. While 16S is targeted to bacteria and reveals their complete spectrum, whole genome sequencing (WGS) enables comparison of metagenomic data at a functional gene level while providing species-level resolution of well-referenced bacteria. Using the same extracted DNA as for 16S analysis, we will perform WGS on the NovaSeq 6000 platform at a depth of 3Gb/sample using the 2×150 bp paired-end protocol. Taxonomy assignment will be determined using MetaPhlAn2 which generates normalized relative abundances/sample/taxon using known marker genes.
- *Bioinformatics:* Functional gene and metabolic pathway (metacyc) profiles will be generated, assigning functional content at both the metagenome and individual bacteria level. Preliminary differences in community and functional profiles according to clinical metadata will be performed in R using non-parametric methods. Bioinformatics’ analyses will be performed using the Quantitative Insights Into Microbial Ecology (QIIME) pipeline as has been previously described [30]. Briefly, the QIIME pipeline utilizes a multi-software approach to perform quality filtering, operational taxonomic unit (OTU) picking, taxonomic assignment, alpha diversity (bacterial diversity within each sample) and beta diversity (bacterial similarities and differences among the samples) measures. With the QIIME pipeline, sequences will be split into sample groups using identification tags on the primers and low-quality sequences will be removed from the dataset.
- Short chain fatty acids (SCFA), including acetate, butyrate and propionate will be analyzed using standard methods [31, 32]. Briefly, the concentrations of SCFA will be measured using a gas chromatography equipped with a flame ionization detector (Hewlett Packard 5890 Series II). Data will be evaluated using an integrator manual (Hewlett Packard 3396 Series II).

#### Beta cell health, function and glycemia

We will collect and compare the following β-cell and glycemic measures pre/post intervention.

- At each visit, fasting proinsulin/C-peptide (PI:C) will be assessed as a measure of β-cell health. Studies from our group [33, 34] have shown that PI:C both at T1D diagnosis and during the honeymoon period remain significantly elevated compared with controls. Proinsulin will be measured by TECOS proinsulin ELISA. C-peptide will be measured using the TOSOH analyzer. PI:C ratios will be calculated as a molar ratio multiplied by 100% to obtain PI as a percentage of C.
- We will collect 2-hour mixed meal tolerance (MMTT)-derived C-peptide measures. MMTTs will be done using standard protocols [35]. C-peptide measures derived from MMTTs will include fasting, area under the curve (AUC) and the peak C-peptide levels as measures of β-cell function.
- Glycemic Measures. HbA1c will be measured at the start of the study and at week 12. Masked CGM will be placed at the baseline visit and at crossover. CGM data will be reviewed to evaluate total time in hypoglycemia, euglycemia, hyperglycemia, mean amplitude of glycemic excursions, the coefficient of variation of glucose levels and time CGM was in use [36-38]. At the baseline/intervention visit and the crossover visit, a blinded CGM sensor will be inserted in those participants who are not currently using a CGM as part of their routine care. Training will be provided on calibration of the sensor, its use in blinded mode, and sensor insertion. Individuals/caregivers will be trained on use of the commercially-available FreeStyle Libre Pro® CGM at their initial visit. This CGM device provides up to 14 days of continuous glucose data upon download at the time of the next study visit. Participants will be reminded to insert a sensor at home after approximately 14 days and continue using the blinded sensor for an additional 14 days. Participants will use their own home blood glucose meter or (if had been previously using one) CGM for regular blood glucose monitoring. The participant will return after 4 weeks to assess the CGM data. CGM must be used for at least 200 hours (equivalent to 8.3 days out of 14 days).
- Data will also be collected on total daily insulin dose and this will be compared before and after the intervention.

#### Dietary Assessments

Dietary Assessments will be obtained using the Automated Self-Administered 24-Hour (ASA24) dietary assessment tool [39-41], a web-based tool for administering automatically coded, self-administered recalls/food records. This will allow assessment of dietary fiber intake along with other dietary components. Participants will be asked to fill these in for the 24 hrs diet recall prior to each stool collection.

Random, non-fasting c-peptide: C-peptide level will be analyzed using a two site immuno-enzymometeric assay using a Tosoh 2000 auto-analyzer (TOSOH, Biosciences, Inc., South San Francisco, CA).

#### Serum and Plasma for storage

subjects will have the option to consent or deny storage. Samples will be used for future type 1 diabetes research assays.

#### MAIT cell analyses

Peripheral blood mononuclear cells (PBMCs) will be isolated from fresh blood samples by Ficoll-Paque (Leucosep) and stained with specific antibodies to quantify and identify MAIT cells, as has been described [17].

- MAIT cell number and phenotype:
  - Flow cytometry and antibodies. Cells will be stained in PBS containing 5% FCS and 0.1% sodium azide. Several antibodies will be used as has been previously described [17]. According to the number of PBMCs obtained from each patient, surface staining will be performed, and when possible due to the number of PBMCs, intracytoplasmic staining of cytokines and GzB will be analyzed after stimulation with PMA and ionomycin, and then PBMC expression of BCL-2 will be analyzed by intracytoplasmic staining (without stimulation). Data acquisition will be performed with a BD Biosciences LSR-Fortessa cytometer or FACSAria III cytometer for cells and with a Beckman Coulter Gallios.
- MAIT cell function:
  - In vitro cell stimulation. For ligand stimulation of human MAIT cells, 5-OP-RU solution will be obtained after incubation of 1 molar equivalent of 5-A-RU with 2 molar equivalents of methylglyoxal (Sigma-Aldrich). HeLa cervical cancer cells expressing MR1 molecules (mycoplasma free), provided by O.L.54, will be cultured with PBMCs in medium containing RPMI medium supplemented with 10% FCS, to analyze activation of MAIT cells in the presence of the MAIT cell ligand (5-OP-RU). In brief, cells will be plated at final concentration of 1×10^6^ PBMCs per ml and 1×10^6^ HeLa cells per ml (1:1 ratio). In these cultures, MAIT cells will be stimulated overnight by the addition of their agonist ligand (5-OP-RU) at various concentrations (0–5 nmol/l) in the presence or absence of blocking mAb MR1 (26.5). Then, MAIT cells will be stained with the appropriate antibodies [17] and cell-surface activation markers will be analyzed by flow cytometry.
  - Intracellular staining. For staining of human BCL-2, after surface staining, lymphocytes will be resuspended in fixation-permeabilization buffer (Foxp3 staining kit from eBioscience) and incubated at 4 °C in the dark, then cells will be washed with PERM Wash buffer (eBioscience) and labeled with the appropriate mAbs (identified above). For analysis of cytokines and granzyme B in MAIT cells, PBMCs obtained from fresh samples will be analyzed after stimulation for 6 h at 37 °C in RPMI medium supplemented with 10% FCS with PMA (25 ng/ml) and ionomycin (1 μg/ml), in the presence of brefeldin A (10 μg/ml).

### Study Endpoints

The primary outcome assessment will be to assess changes in the gut microbiome profile (including frequency and ratio of gram positive to gram negative bacteria) in response to HAMS-AB. Secondary outcome measures will be changes in SCFA levels and changes in pro-insulin to C-peptide ratios as a marker of β-cell health. We will assess β-cell function by responses to an MMTT. We will also examine glycemic outcomes using CGM data and HbA1c measurements. Finally, we will assess the change in number, phenotype and function of MAIT cells in response to intervention with HAMS-AB.

### Data Collection and Schedule of Events

Table 2 outlines the data/samples to be collected and the schedule of events.

**Table 2:** Data Collection.

| Study Phase | Screening Visit | Baseline- Intervention | Washout | Crossover- Intervention | Follow-Up | Early Termination Visit |
| --- | --- | --- | --- | --- | --- | --- |
| Visit Window | | $\leq 30$ days after Visit 1 | +/- 3 days<br>28 days after Visit 2 | +/- 3 days<br>28 days after Visit 3 | +/- 3 days<br>28 days after Visit 4 | |
| Informed consent/ Assent | X |  |  |  |  |  |
| Inclusion/ Exclusion criteria | X |  |  |  |  |  |
| Historical Data/Demographics | X |  |  |  |  |  |
| Random C-peptide concentration | X |  | X |  |  |  |
| Randomization |  | X |  |  |  |  |
| Pregnancy test for females of childbearing age | X | X | X | X | X | X |
| Current Medical data | X | X | X | X | X | X |
| Pubertal Tanner staging with physical exam | X |  |  |  | X | X |
| Ht/Wt measurement and vitals | X | X | X | X | X | X |
| Supervised CGM insertion (and training, if applicable, on use) |  | X |  | X |  |  |
| C-peptide by 2-hr MMTT |  |  | X |  | X | X |
| HbA1c |  | X |  |  | X | X |
| Blood samples for biomarkers (PI:C and *INS* DNA) |  | X | X | X | X | X |
| Provision of HAMS-AB |  | X |  | X |  |  |
| CGM download |  |  | X |  | X | X |
| Stool samples |  | X | X | X | X | X |
| PBMCs for MAIT cell testing |  | X | X | X | X | X |
| Serum and plasma for storage<sup>^</sup> |  | X | X | X | X | X |
| Adverse event assessment |  | X | X | X | X | X |
| Online Dietary Assessments |  | X | X | X | X | X |
| Verbal Diet recall | X |  |  |  |  |  |
| Sampling of prebiotic | X |  |  |  |  |  |

#### Safety Assessments

Adverse events (AEs) will be presented as the proportion of each event seen. This is considered a minimal risk study with low risk for minor adverse events such as mild gastrointestinal symptoms that could be related to increased dietary fiber intake.

Participant information regarding adverse events and safety of the procedure will be collected throughout the study period, including at baseline, weeks 4, 8 and 12. Throughout the study period, participants will be assessed for safety with questions regarding general well-being as well as specific questions to evaluate for adverse events. Participants will also be questioned about stool form and frequency, gastrointestinal symptoms, fevers or infection.

#### Data Safety Monitoring

The data safety monitoring plan involves a pediatric endocrinologist, to review this study for data for quality, rate of subject recruitment, adverse events, procedures for protecting patient privacy, and any protocol deviations every 6 months following the first subject enrolled. If the review concludes that there are significant safety concerns or quality of the data is unsatisfactory, no further enrollment will occur until these issues have been addressed and any changes to the study approved by the IRB. In addition, an internal auditor has been assigned to review the data entry for quality control.

#### Early Termination of a Study Participant

Treatment will stop and the subject will return for an exit visit if any of the following conditions occur:

- withdrawal of consent
- pregnancy (female participants)
- any medically important event such as a concurrent illness or complications

#### Statistical Considerations

A sample size of 12 with a cross over design (i.e. 12 per arm) is considered adequate to estimate effect sizes for efficacy to aid in designing future trials for responses that are normally distributed [42]. Simple comparisons will be performed using chi-square for categorical variables and t-tests for continuous variables. Random attritions will be replaced. An intention to treat analysis will not be appropriate given the crossover design. Our primary outcome is changes in the gut microbiome profile. Microbial sequence analysis results and relative abundance of bacteria will be assessed and biostatistical will be performed using the Quantitative Insights into Microbial Ecology (QIIME) 1.8.0 software as previously described [30].

## Discussion

In summary, here we outline a study protocol using a specific prebiotic, HAMS-AB, in newly diagnosed youth with T1D. We hypothesize that HAMS-AB will improve the gut microbiome profile in children with T1D, increasing SCFA production and that these changes will be associated with improved β-cell function, β-cell health and overall glycemia. We further speculate that these changes will be associated with changes in MAIT cell number and function as well as cytokine changes. We will conduct a single-center pilot randomized cross-over trial using HAMS-AB in 12 youth with newly-diagnosed T1D. We will use state-of-the-art markers to profile the gut microbiome and asses β-cell stress as well as describe frequency, phenotype and function of MAIT cells. In Aim 1, we will determine the gut microbiome profile and SCFA production in youth with recently-diagnosed T1D in response to HAMS-AB consumption. In Aim 2, we will determine the effect of HAMS-AB on glycemia, β- cell function and health. In Aim 3, we will determine the effect of HAMS-AB on frequency, phenotype and function of MAIT cells, anti- and pro-inflammatory cytokine concentrations as well as link these changes with changes in the microbiome profile, glycemia and β-cell health. We anticipate that successful alteration of the gut microbiome using the prebiotic HAMS-AB will be associated with improvement in overall glycemia and a reduction in glycemic variability in recently diagnosed people with T1D as assessed through HbA1c as well as CGM data. We expect that insulin secretion will improve, thus reducing insulin needs. This will be evident from differences in MMTT-derived measures of insulin secretion and reduced insulin doses. We further expect that measures of β-cell stress will show improvement following the use of HAMS-AB. Finally, we anticipate that those with recent onset T1D will have alterations in their number and function of MAIT cells, similar to what has been published and that these changes will show more complete reversion after the intervention.

The effect of gut microbiome modulation on glycemia has been assessed using both prebiotics (supplements containing substrates for the microbiota) and probiotics (supplements containing microbiota). Animal studies have shown that pre and probiotic use can change the gut microbiome profile, modulate the immune system, and improve glucose tolerance [42, 43]. In persons with type 2 diabetes (T2D), prebiotics improve hemoglobin A1c (HbA1c) and postprandial glycemic excursion as well as reduce insulin resistance [44, 45]. This effect on glycemia is thought to be due to mechanisms that include enhanced production and secretion of glucagon-like peptide 1, an incretin hormone that enhances glucose tolerance through modulation of insulin secretion [46]. Altering the gut microbiome may also attenuate a pro-inflammatory milieu, thereby improving insulin sensitivity [47]. Thus, it appears that altering the gut microbiome may improve glycemia, insulin secretion and sensitivity. However, a study using HAMS-AB in newly diagnosed adolescents with T1D has not yet been done.

This is the first study using HAMS-AB in newly diagnosed youth with T1D and has several strengths. If successful, this study will demonstrate that the use of HAMS-AB will be associated with a shift in the gut microbiome to microbial populations capable of HAMS-AB fermentation and increased SCFA production. This is significant since the gut microbiome balance will be restored in these individuals along with increased SCFA levels, known to be associated with increased general health and reduced body inflammation. Further, if we are able to demonstrate an association between consumption of HAMS-AB, improved glycemia and restoration of β-cell function and health, then we may be able to improve diabetes management through the use of a simple inexpensive and safe dietary supplement along with reduced insulin doses used in these patient. In addition, this would serve as the basis to explore using HAMS-AB in those at risk for T1D in order to prevent or delay development of disease. Finally, if indeed are able to show a link between the gut microbiome changes and MAIT cell responses in response to HAMS-AB use, this may lead us to the path to uncover the direct link between immune cell activation and microbiome alterations.

This study has a few limitations. It is a pilot study and therefore, results may not be entirely generalizable. However, data collected from this pilot will allow for larger studies to further explore the findings. In addition, there are several confounders that can affect the gut microbiome. However, by allowing individuals to be their own controls in the study with the current cross-over design, we will minimize the effect of potential confounders such as diet, exercise and environment.

## Data Availability

The data will be generated during the study. Requests for sharing of available data and samples will be granted on a case by case basis.

## Ethics and Dissemination

The study was approved by the Institutional Review Board at Indiana University. Verbal and written information regarding informed consent will be presented to the caregivers and/or patients. Any modifications to the protocol that may affect the conduct of the study will be presented to the committee. The full protocol will be available freely due to open access publication. The findings of this trial will be submitted to a peer-reviewed journal. Abstracts will be submitted to relevant national and international conferences. The standards from the guidelines of the Consolidated Standards of Reporting Trials will be followed for this RCT. All investigators will have access to the final trial dataset.

## Funding and Acknowledgements

We acknowledge the support from the National Institutes of Health, National Center for Advancing Translational Sciences, Clinical and Translational Sciences Award, Grant Numbers, KL2TR002530 (A Carroll, PI), and UL1TR002529 (A. Shekhar, PI). We also acknowledge support from the IU Health Values grant and the Indiana CTSI Project Development Team.

This study design has been previously presented at the Indiana CTSI annual meetings in 2019 and 2020 as well as the Translational Science 2020 meeting.

## Contribution statement

HMI conceived the study, drafted and edited the manuscript. CEM, LAD and MS contributed to the study design, critically reviewed the manuscript and approved the final version.

